# Estimation of causal effects of a time-varying exposure at multiple time points through Multivariable Mendelian randomization

**DOI:** 10.1101/2022.01.04.22268740

**Authors:** Eleanor Sanderson, Tom G Richardson, Tim T Morris, Kate Tilling, George Davey Smith

**Affiliations:** MRC Integrative Epidemiology Unit at the University of Bristol; Population Health Sciences, Bristol Medical School, University of Bristol; Novo Nordisk Research Centre, Headington, Oxford, OX3 7FZ, United Kingdom

## Abstract

Mendelian Randomisation (MR) is a powerful tool in epidemiology to estimate the causal effect of an exposure on an outcome in the presence of unobserved confounding, by utilising genetic variants as instrumental variables (IVs) for the exposure. The effects obtained from MR studies are often interpreted as the lifetime effect of the exposure in question. However, the causal effects of many exposures are thought to vary throughout an individual’s lifetime and there may be periods during which an exposure has more of an effect on a particular outcome. Multivariable MR (MVMR) is an extension of MR that allows for multiple, potentially highly related, exposures to be included in an MR estimation. MVMR estimates the direct effect of each exposure on the outcome conditional on all of the other exposures included in the estimation. We explore the use of MVMR to estimate the direct effect of a single exposure at different time points in an individual’s lifetime on an outcome. We use simulations to illustrate the interpretation of the results from such analyses and the key assumptions required. We show that causal effects at different time periods can be estimated through MVMR when the association between the genetic variants used as instruments and the exposure measured at those time periods varies, however this estimation will not necessarily identify exact time periods over which an exposure has the most effect on the outcome. We illustrate the method through estimation of the causal effects of childhood and adult BMI on smoking behaviour.

## Introduction

Mendelian Randomization (MR) uses the special properties of germline genetic variation to strengthen causal inference of modifiable exposures on disease.[1] MR can be implemented as a form of instrumental variable (IV) estimation that uses genetic variants to estimate causal effects of an exposure on an outcome that is free from bias due to unmeasured confounding.[2–4] As genetic variants which do not change during an individual’s lifetime are used as instruments the estimated effects are interpreted as the lifetime effect of the genetically predicted exposure, or genetic liability for an exposure if that exposure is binary.[5]

Many exposures, such as BMI, may have varying effects on any particular outcome over the course of an individual’s lifetime. Higher BMI in childhood is observationally associated with many health outcomes later in life. However whether this is due to a direct causal effect of childhood BMI on those outcomes or the high correlation between childhood and adult BMI, which then has a causal effect on the outcome, is unclear.[6–9] If a time-varying exposure only affects a (non time-varying) outcome during a particular period then intervening on the exposure during other periods will not have any effect on the outcome. Many disease outcomes occur later in life and understanding if the exposures that are known to associate with the disease have effects over the whole lifecourse or only short-term effects is important for lowering the risk for that disease and understanding individual’s lifetime trajectories. Observational studies often use a lifecourse approach to determine the particular periods in life that affect an outcome[10]. For example, observational studies have shown that sunlight (and from this inferred vitamin D level) in childhood, but not adulthood, is associated with risk of multiple sclerosis.[11–13] Therefore, in order to prevent multiple sclerosis it is important to focus on time spent outside during childhood, intervening in this way during adulthood will not have any effect on the incidence of multiple sclerosis. A lifecourse approach can help to unpick crucial periods for the exposure over an individual’s lifetime which contribute more than others to the outcome in question. This contrasts with a MR approach which will generally provide evidence of a causal effect of the exposure on the outcome regardless of when in the lifecourse the exposure is measured.[14] For example, a MR study has shown a causal effect of vitamin D levels in the aetiology of multiple sclerosis[15] but have not identified which period is important.

When genetic risk for an exposure varies over time MR estimates can be interpreted as the genetically predicted effect of having a higher level of the exposure at the time it is measured.[16] If the genetic variants have a (proportionally) constant effect across the entire lifetime this will be the genetically predicted lifetime effect of having a unit higher level of the exposure across the lifecourse.[17] However, if the genetic variants have differing effects on the exposure at different ages, potentially including no effect at some ages, this effect will be the lifetime effect of having a unit higher level of the exposure at the point it is measured.[16] This will include the effect of the exposure at other points in the lifecourse on the outcome. This may include changes in the level of the exposure of differing magnitudes at different points on the lifecourse. That is; the effect estimated will be the effect on the outcome of having a genetic liability for the exposure that results in a one unit higher genetically predicted level of the exposure at the time the exposure is measured. This allows for the effect of the genetic variants on the exposure to vary over time while still enabling estimation of the causal effect of an exposure that varies over time but which may only be measured at one point in time.

In this paper we explore the use of multivariable Mendelian randomization (MVMR) to estimate the causal effect of a single exposure measured at different time points in an individual’s lifetime on an outcome measured at a single fixed point in time. MVMR is an extension of MR that can be used to estimate the direct effects of multiple, potentially highly related, exposures.[18] Structural mean models have previously been proposed for estimation of MR models with a time varying exposure.[19, 20] The interpretation of the results from estimation of structural mean models will depend on the availability of data for the time-varying exposure, particularly how many points of data are available.[19] MVMR can be implemented when multiple measures of the exposures at different points in the lifecourse are available but can be used to estimate direct effects of the exposure at each of those time points. We outline a model for MR with an exposure measured at multiple time points and explain how this can be estimated with MVMR. We consider specific examples where the assumptions of this model do not hold and present simulation results to investigate what happens in these settings. From these results we explain how the results of such a MVMR estimation can be interpreted. We illustrate these results with application to estimating the effect of child and adulthood BMI on smoking behaviour. The results presented here show that it is possible to estimate genetically predicted direct causal effects of different time periods of an exposure on an outcome using MVMR, however careful interpretation of any results obtained from such an analysis is required.

## Methods

We consider a model where genetic variants are associated with an unmeasured genetic liability for the exposure of interest which in turn affects the observed value of the trait. The liability may differ in different periods of an individual’s lifetime, however the observed trait is likely to change on a more frequent basis reflecting more short term variation or measurement error. Each liability is determined by only the genetic effects and therefore earlier liability levels do not have effects on later liability levels, although they are correlated through shared genetic influences. The observed exposures are influenced by the underlying genetic liability as well as confounders and other environmental influences. These observed exposures have a direct effect on the outcome of interest as well an effect on the subsequent value of the exposure in following periods. The genetic variants used as IV’s are associated with liability in at least one period of the lifecourse but do not have to have the same association with liability in different periods. This allows for the association between a genetic variant and an exposure to vary over different ages, illustrated in Fig 1. This also implies that there may be correlation between the genetic effects during each period. This means that earlier time periods have a total effect on the outcome that is not necessarily equal to their direct effect. This model is given in Fig. 1. IV estimation can correct for the bias introduced by measurement error in the exposure under the assumption that the instrument is not associated with the level of that measurement error.[21] We therefore assume that the exposures are measured without error. All of the variables included (other than the individual genetic variants) are assumed to be continuous and for simplicity we initially limit the number of latent periods to two, however this model could be generalised to any number of periods.

**Figure 1.**
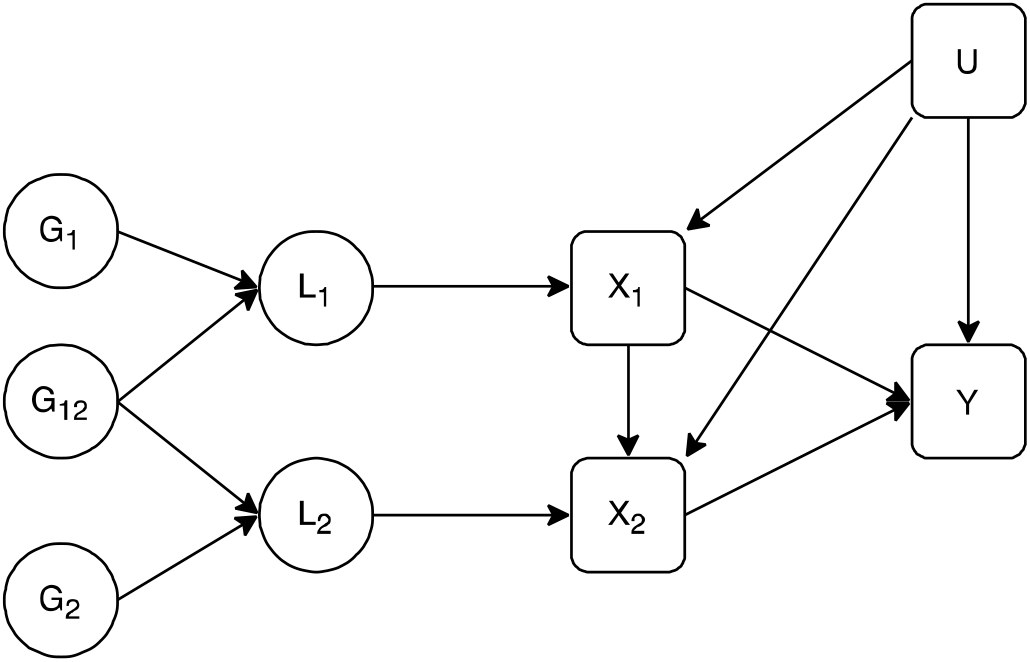
Latent exposure model with two periods of exposure. *L*_1_ is the earlier liability, *L*_2_ is the later liability,*G*_1_ is a set of genetic variants associated with *L*_1_, *G*_2_ is a set of genetic variants associated with *L*_2_, *G*_12_ is a set of genetic variants associated with both *L*_1_ and *L*_2_. *X*_1_ is a measure of the exposure in the early time period *X*_2_ is a measure the exposure at the second time period, *Y* is an outcome observed at one time only, *U* is a set of unobserved confounders of the exposure at each time period and the outcome. *X*_1_ and *X*_2_ are potentially measured with error, error in this measurement is uncorrelated with the genetic variants.

### Estimation with MVMR

MVMR can be used to estimate the genetically predicted effect of the exposure during each period, given the genetically predicted effect of the exposure at all of the other time points included in the estimation, i.e. the effect of *L*_l_ and *L*_2_ in Fig. 1. MVMR can be conducted with either individual level data or summary data and so it is possible to use the methods described here with either type of data. Estimation using individual level data requires a dataset that measured the exposure at all time points considered and the outcome. Summary data from another non-overlapping dataset is required to enable selection of SNPs for use as instruments, conventionally those which are genome-wide significant in a GWAS study. Using summary data requires SNP-exposure effects and SNP-outcome effects taken from separate samples. SNP-exposure associations for the different time points can be taken from either the same or different datasets. Analysis using summary level data is more likely to be feasible in many cases, given the large datasets required and multiple observations at different time points, so we focus here on summary level analysis.

MVMR can be implemented in a summary data setting using estimates of the association between each SNP and: the outcome, 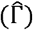; exposure at one time point 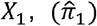; and the exposure at another time point 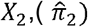, by fitting the following model:

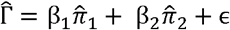

This is a straightforward generalization of the IVW estimation framework for MR.[18, 22]

MVMR estimation relies on three assumptions for estimating the presence or absence of a causal effect of each exposure on the outcome.[18] These assumptions mirror the standard assumptions required for IV estimation and are that; 1. each exposure is robustly predicted by the genetic variants conditional on the other exposures included in the estimation, 2. there is no confounding of the genetic variants and the outcome and 3. the genetic variants are not associated with the outcome other than via exposures included in the estimation, i.e. there are no horizontal pleiotropic effects of the genetic variants on the outcome via other phenotypes. We address the potential for the genetic variants to affect the outcome through the exposure phenotype at time points not included in the estimation in our simulation results.

For multiple exposures to be included in a MVMR the genetic variants must have different effects on each exposure included in the estimation and these effects must not be a linear function of the others.[18] In the model described in Fig. 1, this means the exposure at each time period included in the estimation is associated with a different liability. Whether the genetic variants have differing effects can be tested with a conditional F-statistic.[23, 24] As well as having an F-statistic at each time point greater than 10 to indicate that the genetic variants are strongly associated with that exposure, it is necessary for the conditional F-statistics to be greater than 10, indicating that the genetic variants are robustly associated with exposure at each time period conditional on their association with exposure at the other time period. A large conditional F-statistic indicates that there is enough variation in the genetic effect on the exposure at each time period to identify differences between them.

A heterogeneity Q-statistic can be used to test for violations of the third IV assumption in the MVMR estimation.[18, 23] One potential reason for excessive heterogeneity is that some of the SNPs may be associated with the outcome through pathways that are not included in the MVMR estimation, i.e. there is horizontal pleiotropy. This pleiotropy will bias the results obtained from inverse variance weighted MVMR estimation.[25, 26] If pleiotropy is suspected, alternative estimation methods can be used to estimate MVMR causal effects under different assumptions of the form the pleiotropy takes.[23, 26, 27]

All IV estimation requires additional assumptions for interpretation of the point estimates obtained as causal effects. Firstly, all of the MR and MVMR methods implemented here assume that the causal effects of the exposure(s) on the outcome are linear and, for MVMR, that there are no interactions between the effects of the exposures. Secondly, a ‘point-identifying’ assumption is required. Common point identifying assumptions for univariable IV estimation include homogeneity and monotonicity. The exact definition of this point identifying assumption will determine the precise causal effect estimated, however, it is not currently well-understood how these assumptions relate to estimation with multiple exposures as in MVMR.

### Simulations

#### Inclusion of exposures associated with different latent periods

In the model described in Fig. 1 the genetic variants are associated with the latent variable but not the measured exposure directly. Any measured exposures associated with a common latent period will be associated with the genetic variants in the same way. This means it will not be possible to separately estimate the effects of an exposure at multiple time points which are associated with the same liability. Whether the genetic variants are sufficiently differently associated with each measured time point for the exposure for estimation of the model can be measured with a conditional F-statistic[23, 24], in the same way as MVMR with different exposures.

We illustrate the point above with a simulation. We have included an exposure measured at two time points, both measures of the exposure have a direct causal effect on the outcome and the exposure at the earlier time point also has a small direct effect on the exposure at the later time point. Following the latent exposure model described in Fig. 1 we consider two different structures for the relationship between the genetic variants, the latent exposure and the observed value of the exposure. In the first setting each observed exposure is associated with a different underlying latent period and the genetic variants have different (but correlated) effects on the two latent periods. In the second setting both the observed exposures are associated with the same underlying latent period. This means that the genetic variants have the same effect on the exposure at each time point. These models are illustrated in Fig. 2.

**Figure 2.**
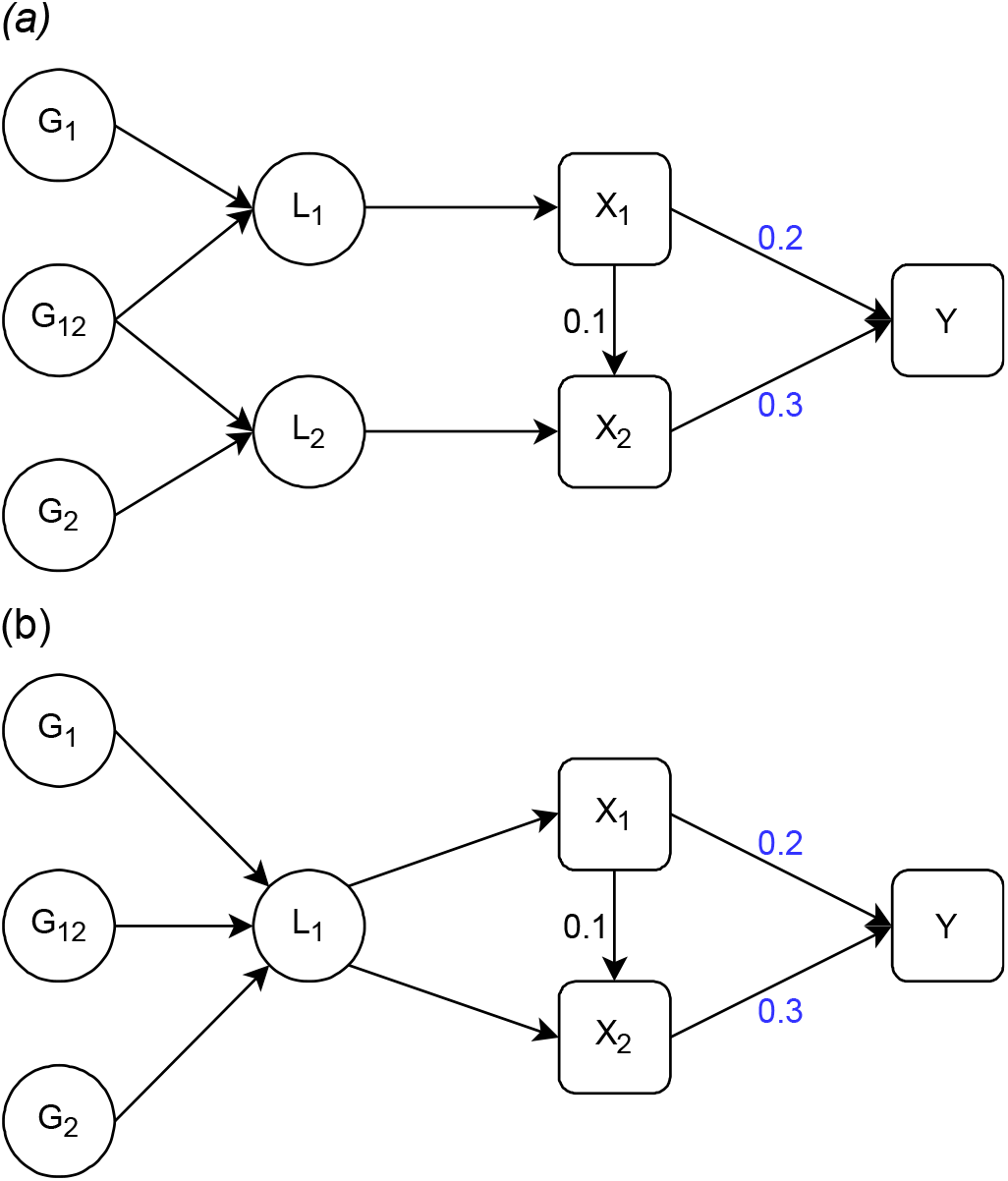
Models with different relationships between the genetic variants and the exposure at each time point. *L*_1_ is the liability in the first time period, *L*_2_ is the liability in the second period. *G*_1_ is a set of genetic variants associated with *L*_1_, *G*_2_ is a set of genetic variants associated with *L*_2_ and *G*_12_ is a set of genetic variants associated with both *L*_1_ and *L*_2_. *X*_1_ and *X*_2_ are observed values of the exposure, where *X*_2_ is observed at a later point in an individual’s life than *X*_1_. *Y* is an outcome. *X*_1_, *X*_2_ and *Y* are confounded by a set of unobserved confounders *U*. In (a) *X*_1_ and *X*_2_ are associated with different liabilities. In (b) *X*_1_ and *X*_2_ are associated with the same liability. *X*_1_ and *X*_2_ are measured with error, though this measurement error is uncorrelated with the genetic variants. The direct causal effect of *X*_1_ on *X*_2_ and *Y*, and *X*_2_ on *Y* are given on the digram.

In each simulation we included 150 SNPs, 2 measures of the continuous exposures and a single continuous outcome. Unobserved confounding was modelled as two continuous variables that affected the earlier exposure measurement and the outcome or the later exposure measurement and the outcome and were excluded from the estimation. These confounders were highly correlated (rho = 0.8). The data simulated were used to generate summary associations between the SNPs and each exposure and the outcome using separate samples, drawn from the same population, for the exposure and the outcome. The true association between the SNPs and each latent period was normally distributed around 0 with variance 0.1/*l* where *l* is the number of SNPs. Effects of the SNPs on each latent period were correlated with *ρ* = 0.25. SNPs associated with the exposure of interest for the MR estimation, or either exposure for the MVMR, with p-values < 5 × 10^−8^ in the exposure sample where included in the estimation. Effect estimates were obtained through inverse variance weighting MVMR (IVW–MVMR).[22] The simulations had a sample size of 150,000 and 2000 repetitions.

Results for the model with either one or two underlying latent periods are given in Table 1. These results show that the univariable estimates give an estimate of the total effect of being on a trajectory that is associated with having a unit higher level of the exposure at the time point associated with the measured exposure. This is larger than either the direct or total effect of the exposure at either time point on the outcome (given in Fig. 2), due to the correlation between the genetic effects on the exposure at each time period. For example, for the first simulation given in Table 1 the direct effect of *X*_l_ on *Y* is 0.20, the total effect is 0.23 and the genetically predicted total effect is 0.34, due to an additional effect of the genetic variants on *X*_2_ which then has an effect on *Y*.

**Table 1.** Simulation results under different relationships between the genetic variants and the exposure at each time point.

|  |  | MR | MVMR |
| --- | --- | --- | --- |
| **Exposures associated with different latent periods** |  |  |  |
| $\beta_1$ | **Genetically predicted lifetime effect** | 0.344 | 0.200 |
|  | Effect estimate | 0.340 | 0.196 |
|  | Est. Std. Error | 0.029 | 0.011 |
|  | Simulation Std. Error | 0.011 | 0.011 |
|  | Absolute bias | 0.010 | 0.009 |
|  | F-statistic | 96.313 |  |
|  | Conditional F-statistic |  | 55.758 |
| $\beta_2$ | **Genetically predicted lifetime effect** | 0.376 | 0.300 |
|  | Effect estimate | 0.371 | 0.297 |
|  | Est. Std. Error | 0.015 | 0.009 |
|  | Simulation Std. Error | 0.008 | 0.009 |
|  | Absolute bias | 0.008 | 0.008 |
|  | F-statistic | 129.308 |  |
|  | Conditional F-statistic |  | 78.006 |
| **Exposures associated with the same latent period** |  |  |  |
| $\beta_1$ | **Genetically predicted lifetime effect** | 0.530 | 0.200 |
|  | Effect estimate | 0.519 | 0.207 |
|  | Est. Std. Error | 0.011 | 0.080 |
|  | Simulation Std. Error | 0.011 | 0.080 |
|  | Absolute bias | 0.013 | 0.063 |
|  | F-statistic | 96.313 |  |
|  | Conditional F-statistic |  | 1.059 |
| $\beta_2$ | **Genetically predicted lifetime effect** | 0.480 | 0.300 |
|  | Effect estimate | 0.474 | 0.288 |
|  | Est. Std. Error | 0.009 | 0.073 |
|  | Simulation Std. Error | 0.009 | 0.072 |
|  | Absolute bias | 0.009 | 0.058 |
|  | F-statistic | 115.758 |  |
|  | Conditional F-statistic |  | 1.062 |

N= 150,000, reps = 2000, $\beta_1 = 0.2$ , $\beta_2 = 0.3$ . Effect of $X_1$ on $X_2 = 0.1$ . True genetically predicted effects for each estimation are given in the table. Absolute bias is the mean value of the absolute bias of the effect estimate across the simulations. For each of *Effect estimate*, *Est. Std Error*, *F-statistic* and *Conditional F-statistic* mean values across each iteration of the simulation are reported. *Simulation Std. Error* is the estimated standard error in the effect estimate across the repetitions in the simulation.

When the measured exposures are associated with different latent periods, MVMR consistently estimates the genetically predicted causal effect of being on a trajectory associated with a unit higher level of that exposure, given the level of the exposure at the other time period. However, when the measured exposures are associated with the same latent period there is no difference in the genetic effects on the measured exposures and therefore weak instrument bias is introduced into the MVMR estimation.[23] This is highlighted through low conditional F-statistics. In this setting there is random variation in the direction of the bias for each exposure in each repetition of the simulation. Therefore the mean point estimate is close to the true value of the causal effect. However, the high mean level of absolute bias shows the bias from conditionally weak instruments. This highlights how the MVMR estimates are not only biased by weak instruments but that the bias could act in either direction, with different repetitions within the same simulation being biased in opposite directions.

We also explored the effect of only selecting genetic variants which had differing effects at each time point on the results obtained for each of the models described here, as has previously been applied elsewhere.[28] This analysis shows that although this causes apparent conditional instrument strength to increase the causal effect estimates are potentially biased due to genetic variants which differ in the effects on each exposure more than others by chance by being selected for the analysis. These estimation results also have lower power than those using all SNPs due to the reduction in the number of genetic variants included. We therefore recommend that this approach is avoided and do not consider it further. Results from this estimation and a full description of the analysis are given in the Supplementary material.

#### Estimation in the presence of a causal effect from the outcome to the later time point

We now consider a model where the outcome has an effect on the exposure measured at the later time point. The exposure at the later time period is therefore a collider of the earlier exposure and the outcome. This is illustrated in Fig 3 and in all other aspects the model is the same as that described in Fig 2(a). Morris et al (2021) showed that estimation of this scenario with MR gives consistent estimates when there is a single underlying latent period.[16] Here we consider MVMR estimation of a model with two underlying latent periods.

**Figure 3.**
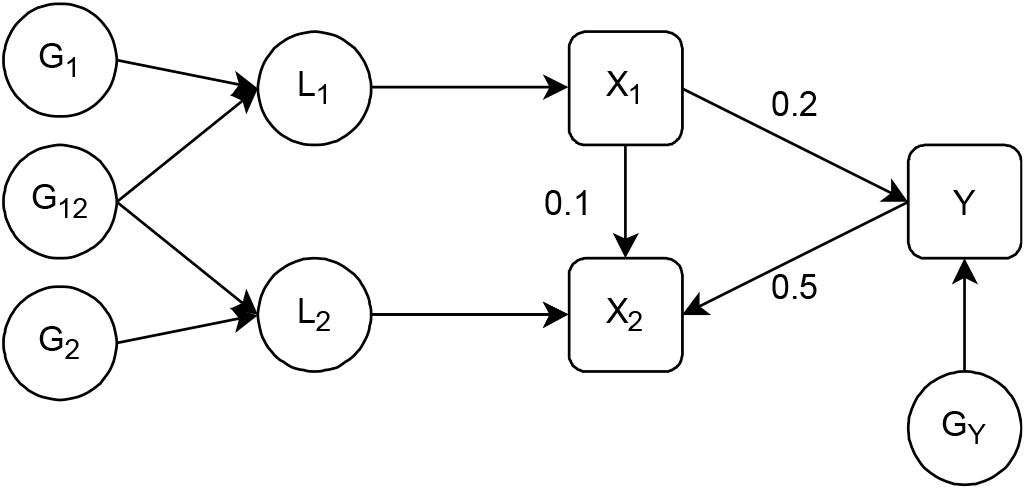
Model with a causal effect from the outcome to the later time point. *L*_1_ is the liability in the first time period, *L*_2_ is the liability in the second period. *G*_1_ is a set of genetic variants associated with *L*_1_, *G*_2_ is a set of genetic variants associated with *L*_2_ and *G*_12_ is a set of genetic variants associated with both *L*_1_ and *L*_2_. *G*_*Y*_ is a set of genetic variants associated with the outcome. *X*_1_ and *X*_2_ are observed values of the exposure, where *X*_2_ is observed at a later point in an individual’s life than *X*_1_. *Y* is an outcome. *X*_1_, *X*_2_ and *Y* are confounded by a set of unobserved confounders *U. X*_1_ and *X*_2_ are measured with error, this measurement error is uncorrelated with the genetic variants. The direct causal effect of *X*_1_ on *X*_2_ and *Y*, and *Y* on *X*_2_ are given on the digram.

Simulations were set up in the same way as described for Table 1 with the addition of 50 SNPs included that were associated with the outcome *Y*. This model was estimated assuming that *X*_l_ and *X*_2_ are the true exposures and *Y* is the true outcome. As all genetic variants associated with the exposure at either time period, selected based on a p-value for the SNP – exposure association of <5×10^−8^, reflecting genome-wide significance, were included in the MVMR estimation some SNP strongly associated with *Y* were selected as instruments for the later time period. Results from this simulation are given in Table 2.

**Table 2.**
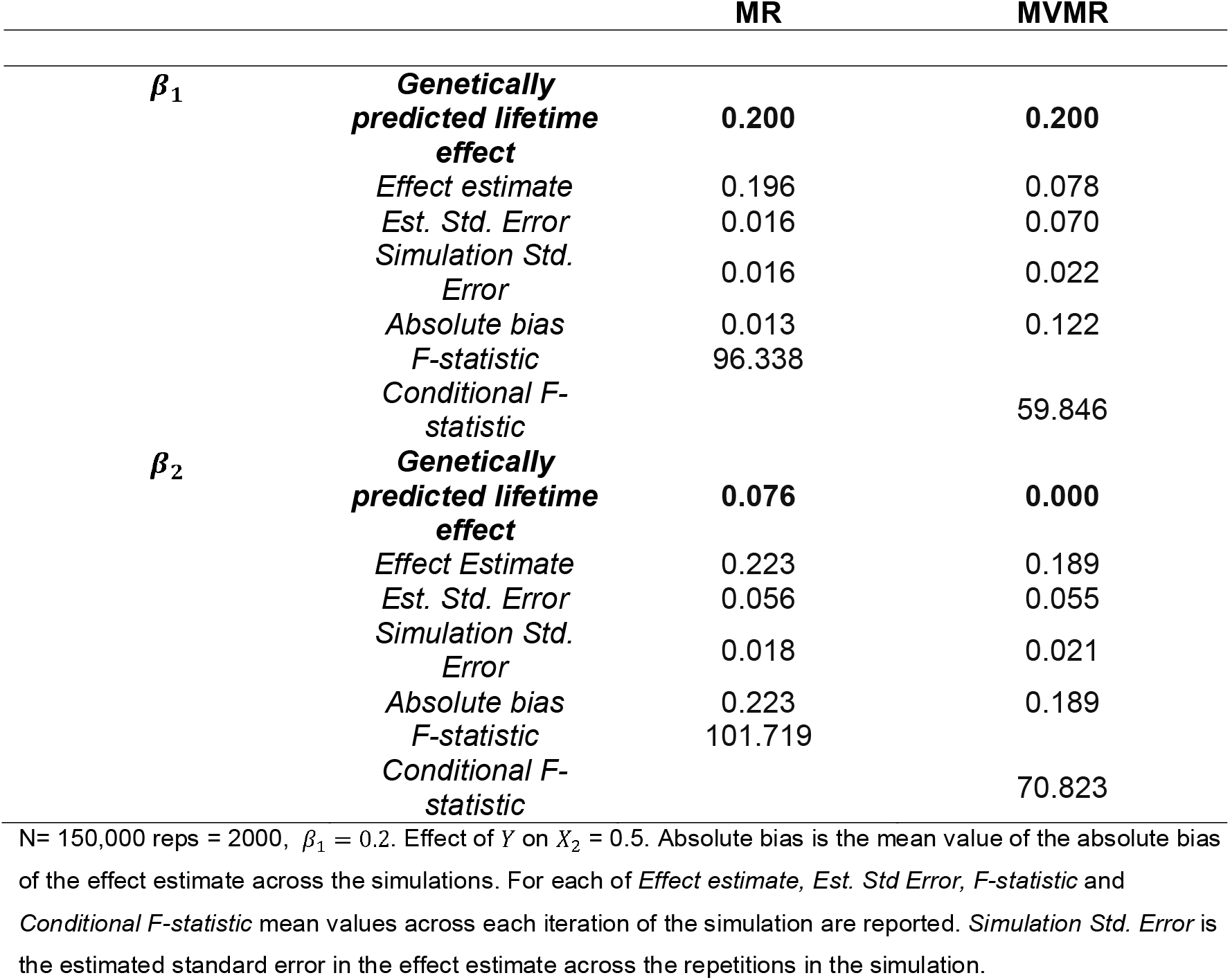
Simulation results for multiple time points with a causal effect from the outcome to the later time point.

These simulation results show that although the genetic variants strongly predict the exposure at each time period conditional on the other, MVMR estimation gives biased estimates of the direct causal effect of the exposure at both time periods on *Y*. This bias is due to conditioning on a variable that depends on both the exposure and the outcome (a collider) in the estimation, introducing collider bias.[29–32] Because the genetically predicted value of *X*_2_ depends on genetic variants associated with *Y, X*_2_ becomes the collider in the MVMR estimation. Conditioning on a collider distorts the estimated association between the other exposure and the outcome and so means that the estimates obtained in the MVMR are no longer reliable estimates of the direct effect of the earlier exposure on the outcome. Sanderson et al. (2019) showed that MVMR conditioning on a collider does not introduce collider bias when only genetic variants associated with the exposures are included in the estimation.[18] The different result here occurs because we have allowed for genetic variants associated with Y to be included as instruments, which was not the case in Sanderson et al. (2019) and reflects a situation where the primary phenotype has been mis-specified.[2] This collider bias can be avoided if the genetic variants used in the estimation are restricted to those which affect the exposure at either time point directly without acting via the outcome, however identifying these variants is not always easy. Importantly, the introduction of collider bias in this estimation biases the effect estimates at each time point included in the estimation, including the earlier time point which is not dependent on *Y*.

#### Additional excluded latent time period

We finally consider a model where the exposure has three underlying latent periods associated with it but where the model estimated only includes the exposure at times associated with two of those latent periods. The true structure of the data is illustrated in Fig. 4 however the model estimated is assumed to be the same as that given in Fig 2(a).

**Figure 4.**
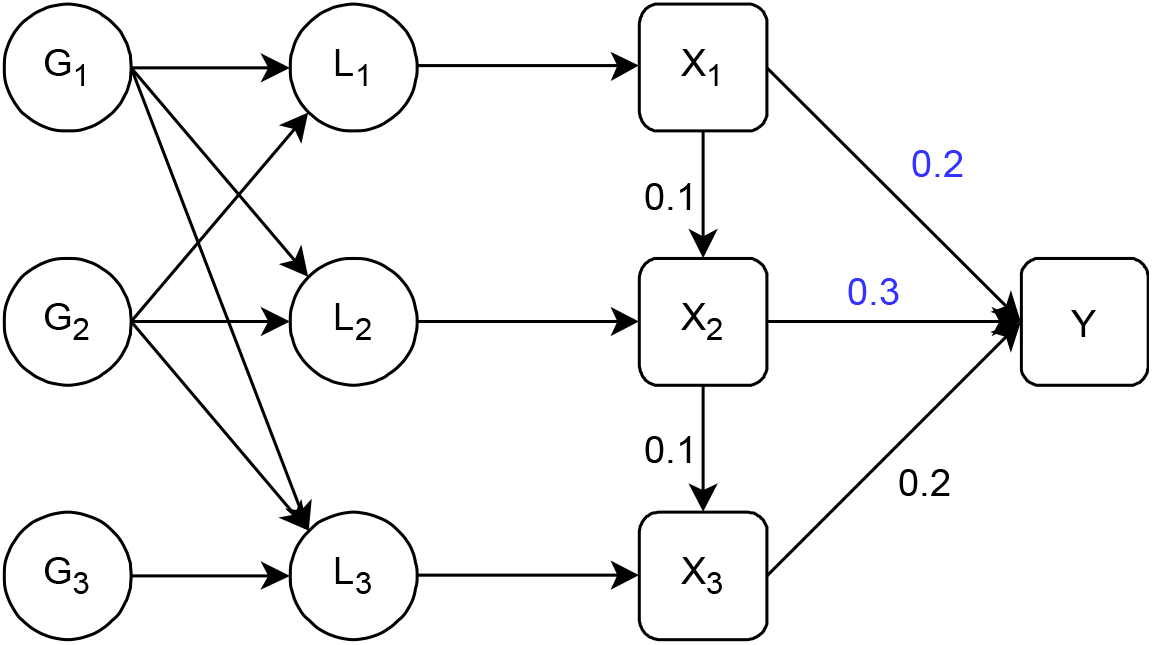
Model with three latent time periods. *L*_1_ is the latent value of the exposure in the first time period, *L*_2_ is the latent value of the exposure in the second period, *L*_3_ is the latent value of the exposure in the third period, *G*_1_ is a set of genetic variants associated with *L*_1_, *L*_2_ and *L*_3_, *G*_2_ is another set of genetic variants associated with *L*_1_, *L*_2_ and *L*_3_ and *G*_3_ is a set of genetic variants associated with *L*_3_. *X*_1_ and *X*_2_ are observed values of the exposure, where *X*_2_ is observed at a later point in an individual’s life than *X*_1_. *X*_3_ is a third value of the exposure that is not observed in our simulation. *Y* is an outcome. *X*_1_, *X*_2_, *X*_3_ and *Y* are confounded by a set of unobserved confounders *U. X*_1_, *X*_2_ and *X*_3_ are measured with error, this measurement error is uncorrelated with the genetic variants. The direct causal effect of *X*_1_ on *X*_2_ and *Y, X*_2_ on *X*_3_ and *Y*, and *X*_3_ on *Y* are given on the digram.

We set the simulations up in the same way as described for Table 1 with the addition of a third latent time period associated with a measured value of the exposure. This third measured exposure is assumed to be dependent on *X*_2_ and subject to overlapping confounding to both *X*_l_ and *X*_2_ with *Y*. We considered two models for the effect of *G* on *L*_3_. In the first there is no correlation between the association between *G* and *L*_3_ and the association between *G* and the other liabilities. In the second correlation between the association between *G* and *L*_3_ and *G* and *L*_l_ and *L*_2_ was added with higher correlation between *G* − *L*_2_ and *G* − *L*_3_ (*ρ* = 0.25) and a lower level of correlation between *G* − *L*_l_ and *G* − *L*_3_ (*ρ* = 0.1). These correlations arise from the overlap in the genetic effects on each liability. In both cases the outcome is assumed to occur at or after the time at which *X*_3_ is measured and all exposures have a direct causal effect on the outcome. The results from this simulation are given in Table 3.

**Table 3.** Simulation results with a relevant latent period excluded.

|  |  | MR | MVMR |
| --- | --- | --- | --- |
| **Correlated genetic effects** |  |  |  |
| $\beta_1$ | **Genetically predicted lifetime effect** | **0.399** | **0.226** |
|  | Effect estimate | 0.387 | 0.210 |
|  | Est. Std. Error | 0.038 | 0.017 |
|  | Simulation Std. Error | 0.018 | 0.018 |
|  | Absolute bias | 0.015 | 0.017 |
|  | F-statistic | 96.457 |  |
|  | Conditional F-statistic |  | 55.834 |
| $\beta_2$ | **Genetically predicted lifetime effect** | **0.479** | **0.371** |
|  | Effect estimate | 0.446 | 0.366 |
|  | Est. Std. Error | 0.019 | 0.015 |
|  | Simulation Std. Error | 0.014 | 0.015 |
|  | Absolute bias | 0.033 | 0.010 |
|  | F-statistic | 129.482 |  |
|  | Conditional F-statistic |  | 78.068 |
| **Independent genetic effects** |  |  |  |
| $\beta_1$ | **Genetically predicted lifetime effect** | **0.345** | **0.199** |
|  | Effect Estimate | 0.348 | 0.195 |
|  | Est. Std. Error | 0.036 | 0.023 |
|  | Simulation Std. Error | 0.022 | 0.024 |
|  | Absolute bias | 0.011 | 0.011 |
|  | F-statistic | 96.457 |  |
|  | Conditional F-statistic |  | 55.834 |
| $\beta_2$ | **Genetically predicted lifetime effect** | **0.396** | **0.320** |
|  | Effect estimate | 0.391 | 0.317 |
|  | Est. Std. Error | 0.022 | 0.020 |
|  | Simulation Std. Error | 0.018 | 0.020 |
|  | Absolute bias | 0.009 | 0.017 |
|  | F-statistic | 129.482 |  |
|  | Conditional F-statistic |  | 78.068 |

**Table 4.**
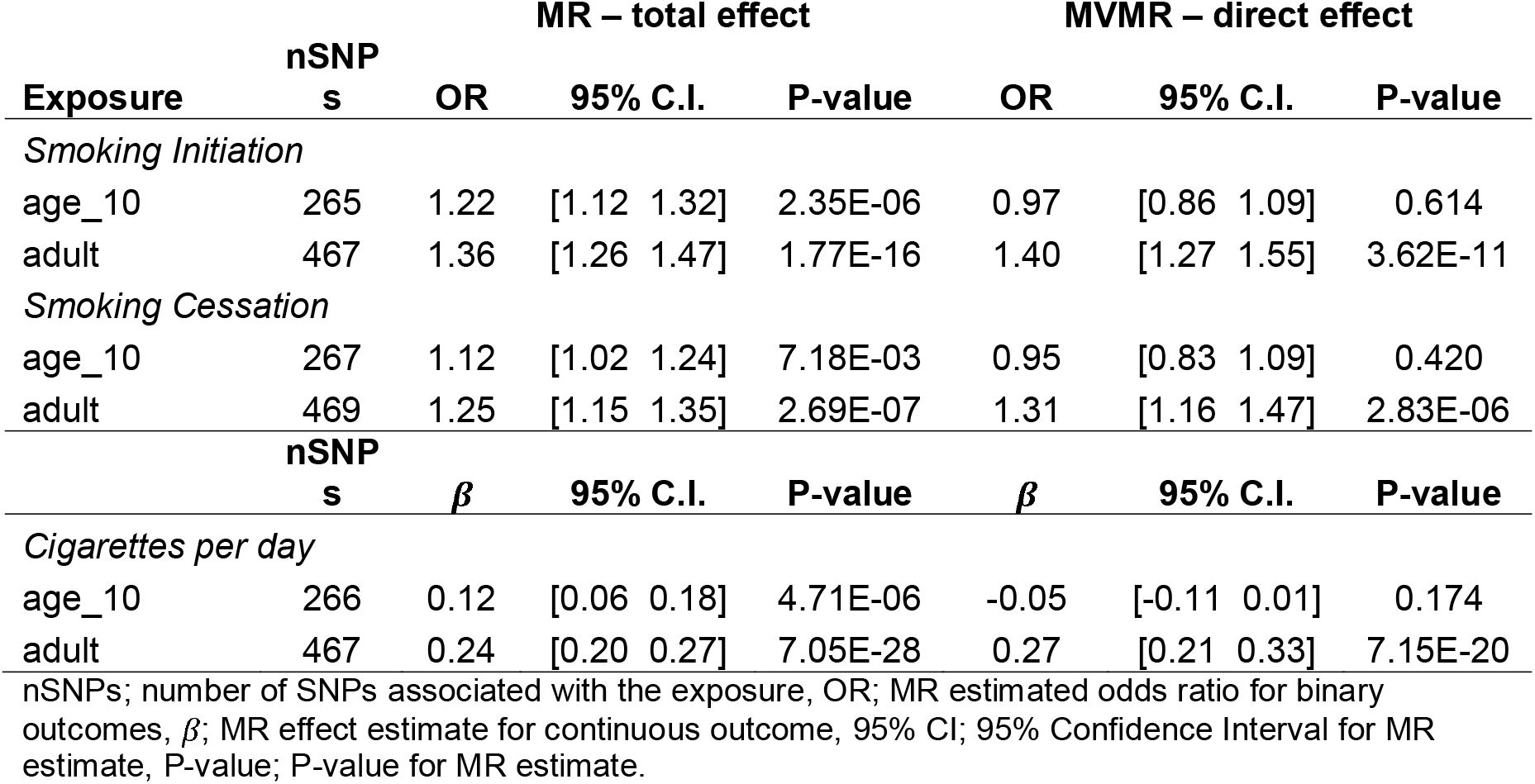
Univariable and multivariable estimates for effect of child and adulthood BMI on smoking behaviour

|  |  | MR – total effect |  |  |  | MVMR – direct effect |  |  |  |
| --- | --- | --- | --- | --- | --- | --- | --- | --- | --- |
|  | nSNP |  |  |  |  |  |  |  |  |
| Exposure | s | OR | 95% C.I. |  | P-value | OR | 95% C.I. |  | P-value |
| Smoking Initiation |  |  |  |  |  |  |  |  |  |
| age_10 | 265 | 1.22 | [1.12 | 1.32] | 2.35E-06 | 0.97 | [0.86 | 1.09] | 0.614 |
| adult | 467 | 1.36 | [1.26 | 1.47] | 1.77E-16 | 1.40 | [1.27 | 1.55] | 3.62E-11 |
| Smoking Cessation |  |  |  |  |  |  |  |  |  |
| age_10 | 267 | 1.12 | [1.02 | 1.24] | 7.18E-03 | 0.95 | [0.83 | 1.09] | 0.420 |
| adult | 469 | 1.25 | [1.15 | 1.35] | 2.69E-07 | 1.31 | [1.16 | 1.47] | 2.83E-06 |
|  | nSNP |  |  |  |  |  |  |  |  |
| | s | $\beta$ | 95% C.I. | | P-value | $\beta$ | 95% C.I. | | P-value |
| Cigarettes per day |  |  |  |  |  |  |  |  |  |
| age_10 | 266 | 0.12 | [0.06 | 0.18] | 4.71E-06 | -0.05 | [-0.11 | 0.01] | 0.174 |
| adult | 467 | 0.24 | [0.20 | 0.27] | 7.05E-28 | 0.27 | [0.21 | 0.33] | 7.15E-20 |

nSNPs; number of SNPs associated with the exposure, OR; MR estimated odds ratio for binary outcomes, $\beta$ ; MR effect estimate for continuous outcome, 95% CI; 95% Confidence Interval for MR estimate, P-value; P-value for MR estimate.

The liability in the first and second periods are correlated due to the overlap in the genetic effects on each liability. When the association between the genetic variants and the excluded time period are correlated with the associations for the included periods the effect estimated will include some of the effect that acts via the omitted time periods. The estimated effect of *X*_2_ therefore differs from the direct effect by a larger amount that the estimated effect of *X*_l_, however each is a consistent estimate of the effect of being on a trajectory associated with a unit higher exposure at that time point. This is due to the particular structure of the data in our simulation, with a larger correlation between the genetic effects on *L*_2_ and *L*_3_ than between the genetic effects on *L*_l_ and *L*_3_ and would not be expected to apply in all cases. When the genetic effects on *L*_3_ are uncorrelated with the included latent periods the effect of being on a unit higher trajectory at that time point is the same as the direct effect. However, in this case the genetically predicted direct effect of *X*_2_ on *Y* is larger than the direct effect (0.2) because this effect includes the effect that acts via the measured value associated with the excluded latent period, *X*_3_.

## Application

We consider an illustrative application where we estimate the effect of childhood and adult body mass index (BMI) on smoking behaviour, measured as smoking initiation, smoking cessation and cigarettes per day.

### Data

Data on child and adulthood BMI were taken from the UK biobank (UKB) study. [33, 34] Between 2006 and 2010, the UK Biobank study enrolled 500,000 individuals aged between 40 and 69 at baseline across 22 assessment centres in the UK. Data were collected on clinical examinations, assays of biological samples, detailed information regarding self-reported health characteristics and genome-wide genotyping. In total 12,370,749 genetic variants in up to 463,005 individuals were available for analysis, as described previously.[35] For BMI we derived a measure of childhood body size using recall questionnaire data asking UKB participants if they were ‘thinner’, ‘plumper’ or ‘about average’ when they were aged 10 years old compared to the average. Adult body size was derived using clinically measured BMI data (mean age 56.5 years), which we categorized into a 3-category variable using the same proportion as the early life measure for comparative purposes. Genetic variants robustly associated with childhood and adult body size (based on P<5×10^−8^ and r^2^<0.001 using a reference panel from the 1000 genomes project phase 3 [36]) were identified from a previously undertaken Genome Wide Association Study (GWAS) in UKB. This GWAS has been described in-detail elsewhere as well as validation studies of the resulting genetic instruments.[37–39]

Each of the smoking behaviour outcomes GWAS data was extracted from Lui et al (2019) using summary statistics produced excluding UKB.[40] The mean age of smoking initiation across individuals with available data (excluding UK Biobank) was 17.5 years, with the mean for each study included in the GWAS ranging from 16.0 to 21.0 years.

For each outcome considered we estimated the genetically predicted total effect of early life and later life exposure separately through a two-sample MR using the SNPs associated with the exposure at the relevant time period. We then estimated the genetically predicted direct effects of the exposure at each time point through a MVMR estimation including both early and later life body size in the same estimation, including all SNPs associated with the exposure at either time.

### Results

Our MR estimates showed a strong total effect of body size in childhood and adulthood on all of the smoking outcomes (total effect of a category increase in childhood body size on number of cigarettes per day = 0.13, 95% CI=0.07 to 0.18, P=2.11×10^−6^, for adult body size: 0.25, 95% CI=0.20 to 0.30, P=3.54×10^−26^). However, in the MVMR no effect of early life body size on number of cigarettes per day was observed and the effect of later life remained largely unchanged implying that the total and direct effects of later life body size are similar (direct effect of a category increase childhood body size on number of cigarettes per day = −0.05, 95% CI=-0.11 to 0.01, P=0.174, for adult body size; 0.27, 95% CI=0.22 to 0.35, P=7.15×10^−20^). Similar results were observed for the other smoking behaviour measures with positive total effects of higher category of childhood body size on smoking initiation and cessation observed in the MR estimation and no direct effect of childhood body size observed in the MVMR estimation. These results suggest that there is no direct effect of childhood body size on smoking behaviour in later life. The observed effect in the MR estimates of childhood body size on smoking are due to a combination of the effect of SNPs associated with childhood body size also having an effect on adult body size and an indirect effect of childhood body size on smoking behaviour through its effect on adult body size. Steiger filtering[41] between adult body size and the outcome removed very few (≤ 5) SNPs for any of the smoking behaviours and did not change the results obtained, results given in Supplementary Table 2.

We have not explored the potential for biases that often arise in MR and MVMR studies in the results presented here, such as biases due to pleiotropy or collider bias.[25, 31, 42] These results should therefore be taken as an illustration of the application and interpretation of the methods discussed.

## Discussion

When multiple measures of an exposure at different time points are available, MVMR can be used to estimate the causal effect of changing the liability of the exposure at different time points on the outcome. The interpretation of the estimate is the direct effect of having a liability associated with a unit higher level of the exposure at that time point, for a given liability for the exposure at the other time points included in the estimation. That is, the effect of having a liability associated with a unit higher level of *X*_l_ while keeping the liability for *X*_2_ constant. If measures of the exposure at different time periods are available, it is possible to identify whether the exposure effects persist over time or key periods exist in the lifecourse.

An important restriction for estimation of these models is that the association between the genetic variants used as instruments and the exposure must vary over the periods included in the estimation. Although genetic variants do not vary over an individual’s lifetime, variation could arise from different genetic variants having different levels of importance in the development of the exposure at different ages. If the genetic variants associated with liability at a particular time are also associated with liability for a period excluded from the estimation, the estimated effect obtained will include that part of the effect of the excluded period. In our simulations we have assumed that each liability only affects the exposure at one time period but that genetic variants can affect multiple liabilities. However, the results obtained would be the same if we had allowed each genetic variant to influence one liability only but for the liabilities to affect the exposure at multiple time periods and each exposure to be influenced by multiple liabilities.

The effects of any time periods excluded from the estimation but associated with genetic variants included in the estimation will form part of the effect estimated. Therefore, if MVMR is used to identify important periods in the lifecourse, then other potentially important periods also need to be included in the estimation. Previous work using this estimation approach has shown that early life BMI does not have a direct effect on type 2 diabetes and coronary heart disease.[37] Therefore if an individual with a high BMI in early life reduces their excess weight in later life their risk for type 2 diabetes and coronary heart disease will not be increased via this pathway. In contrast, our application shows an effect of adult BMI (mean age: 56.5) on smoking initiation (mean age: 17.5 years), once childhood BMI has been controlled for. Typical age of smoking initiation precedes adulthood however, the effect estimated is the effect of being on a trajectory associated with a higher level of BMI in adulthood, rather than the effect at that time point. The large effect of BMI at the age measured in our sample on risk of smoking is therefore unlikely to be causal at the point of time that the exposure and outcome were measured. If higher liability for BMI in adulthood is associated with higher liability for BMI in adolescence the effect estimated may represent the effect of higher liability for BMI in adolescence on smoking initiation even though BMI is measured at a later time point. This model is illustrated in Fig. 5. Data on BMI at different ages between childhood and adulthood would potentially enable estimation of the effect of BMI on smoking behaviour at a range of different times and so identify the period between childhood and adulthood that was most important. Implementation of this approach with MVMR would however rely on those periods being differentially associated with the genetic variants used as instruments.

**Figure 5.**
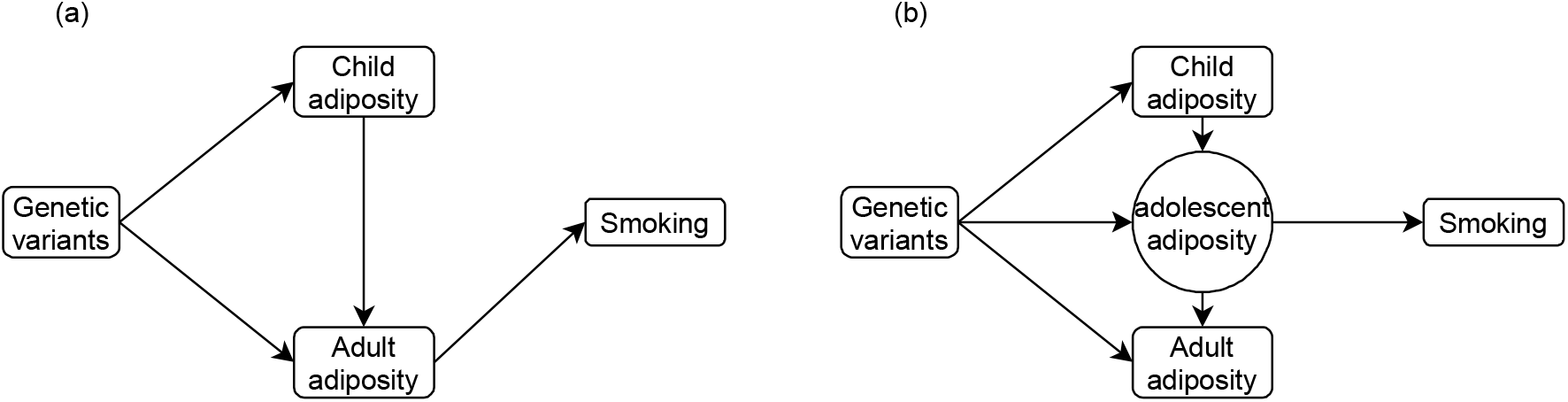
Estimated relationship between adiposity and smoking behavior. Figure shows (a) the estimated relationship between adiposity and smoking behaviour and (b) a potential underlying model that would give this result.

A key assumption for the implementation of this method is that the association between genetic variants and the exposure of interest varies over the lifecourse. This may not be the case for all exposures, however it has been shown for body mass index (BMI) previously. For example; Hardy et al (2010) show that the association between two genetic variants in FTO and near MC4R vary in their association with BMI/body size over different ages using data for individuals aged 2 – 53.[43] Richardson et al (2020) conducted a genome-wide association study of self-reported body size at age 10 and adulthood BMI in UK Biobank and showed that the estimated association for many genetic variants differed between these two ages.[37] Twin studies have also shown that the genetic influence on BMI changes over different ages.[44] However other research has shown that the genetic influences on BMI are consistent across adulthood.[45] This would prevent the estimation of causal effects of different periods within adulthood using MVMR analysis. The assumption that genetic effects differ for each period should be considered for every application and can be tested with a conditional F-statistic.[18, 23]

An assumption that all of the data is from the same underlying population is important to all summary-data MR analysis.[46, 47] This is likely to be particularly important when considering the same exposure at different ages as changes in the distribution of the exposure or the relationship between the exposure and the outcome between different cohorts could potentially bias the results obtained. The choice of datasets should be carefully considered if the same data cannot be used for each time point, for example; the distribution of childhood BMI levels has changed notably over the last 50 years and therefore it would not be correct to assume that BMI measured in a group of adults and children now would represent measures from the same population.

An important assumption of all MR analyses is the assumption of no horizontal pleiotropy, i.e. that the exclusion restriction holds. This can be assessed in MVMR through examination of the Q-statistic for heterogeneity and the application of pleiotropy robust methods.[18, 26, 27] However there may be heterogeneity in the SNP effects even in the absence of pleiotropy if the SNPs are associated with different trajectories for the exposure and the causal effect of the exposure varies over time. In this case those SNPs that have a larger association with the exposure in the time period with the largest causal effect will estimate a larger causal effect of the exposure on the outcome. This will inflate the heterogeneity Q-statistic even in the absence of conventional pleiotropic effects.

Finally, we have throughout this work considered only a single measurement of the outcome. For many exposures and outcomes it may be possible that the outcome could also vary over time with the relationship between the exposure and outcome varying at different time points, and potentially also effects of earlier values of the exposure on later values of the outcome. This sort of relationship, with multiple different outcomes, cannot be estimated with standard MVMR methods. This is therefore left as an area of future research.

## Data Availability

Data used are available through application to UK Biobank or are publicly available summary statistics.

## Ethical approval

UK biobank has received ethical approval from the UK National Health Service’s National Research Ethics Service (re 11/NW/0382) and data were analysed in this project under app #15825. All other data analysed were from publicly available summary statistics generated using relevant ethical approval from their respective studies.

## Code availability

Code for the simulation study is available at https://github.com/eleanorsanderson/Child_AdultBMI.

## Author Contributions

ES conducted the simulation study and wrote the first draft of the manuscript. TGR conducted the applied analysis. All authors reviewed and edited the manuscript.

