## Supplementary Table 1 for "Estimation of causal effects of a time-varying exposure at multiple time points through Multivariable Mendelian randomization"

**SUPPLEMENTARY MATERIAL**

Eleanor Sanderson^1,2^, Tom G Richardson^1,2,3^, Tim T Morris^1,2^, Kate Tilling^1,2^, George Davey Smith^1,2^

1. *MRC Integrative Epidemiology Unit at the University of Bristol.*
2. *Population Health Sciences, Bristol Medical School, University of Bristol.*
3. *Novo Nordisk Research Centre, Headington, Oxford, OX3 7FZ, United Kingdom*

**Supplementary Table 1 –** *Simulation results under different relationships between the genetic variants and the exposure at each time point with selected variants.*

*For the MVMR estimation genetic variants have been selected that have the largest difference in effect on each time period only. These were defined as those in the top 20% for the absolute difference of effect on the exposure in each period.*

|  |  | **MR** | **MVMR** |
| --- | --- | --- | --- |
| ***Exposures associated with different latent periods*** | |  |  |
| $\boldsymbol{\beta}_{\boldsymbol{1}}$ | ***Genetically predicted lifetime effect*** | 0.344 | 0.200 |
|  | *Effect estimate* | 0.340 | 0.196 |
|  | *Est. Std. Error* | 0.029 | 0.021 |
|  | *Simulation Std. Error* | 0.011 | 0.020 |
|  | *Absolute bias* | 0.010 | 0.016 |
|  | *F-statistic* | 96.31 |  |
|  | *Conditional F-statistic* |  | 62.88 |
| $\boldsymbol{\beta}_{\boldsymbol{2}}$ | ***Genetically predicted lifetime effect*** | 0.376 | 0.300 |
|  | *Effect estimate* | 0.371 | 0.297 |
|  | *Est. Std. Error* | 0.015 | 0.016 |
|  | *Simulation Std. Error* | 0.008 | 0.016 |
|  | *Absolute bias* | 0.008 | 0.013 |
|  | *F-statistic* | 129.308 |  |
|  | *Conditional F-statistic* |  | 94.604 |
| ***Exposures associated with the same latent period*** | |  |  |
| $\boldsymbol{\beta}_{\boldsymbol{1}}$ | ***Genetically predicted lifetime effect*** | 0.530 | 0.200 |
|  | *Effect estimate* | 0.519 | 0.196 |
|  | *Est. Std. Error* | 0.011 | 0.175 |
|  | *Simulation Std. Error* | 0.011 | 0.185 |
|  | *Absolute bias* | 0.013 | 0.143 |
|  | *F-statistic* | 96.313 |  |
|  | *Conditional F-statistic* |  | 1.396 |
| $\boldsymbol{\beta}_{\boldsymbol{2}}$ | ***Genetically predicted lifetime effect*** | 0.480 | 0.300 |
|  | *Effect estimate* | 0.474 | 0.296 |
|  | *Est. Std. Error* | 0.009 | 0.141 |
|  | *Simulation Std. Error* | 0.009 | 0.149 |
|  | *Absolute bias* | 0.009 | 0.116 |
|  | *F-statistic* | 115.758 |  |
|  | *Conditional F-statistic* |  | 1.406 |

N= 150,000, reps = 2000, $\beta_{1}=0.2$, $\beta_{2}=0.3$. Effect of X_1_ on X_2_ = 0.1. True genetically predicted effects for each estimation are given in the table. Absolute bias is the mean value of the absolute bias of the effect estimate across the simulations. For each of *Effect estimate, Est. Std Error, F-statistic* and *Conditional F-statistic* mean values across each iteration of the simulation are reported. *Simulation Std. Error* is the estimated standard error in the effect estimate across the repetitions in the simulation.

**Supplementary Table 2 –** *Univariable and multivariable estimates for effect of child and adulthood BMI on smoking behaviour with Steiger filtering applied.*

*Steiger filtering has been applied to remove any SNPs that predict more variation in adult BMI category than in the smoking outcome considered. No SNPs were removed for smoking initiation, 1 SNP was removed for smoking cessation and 5 SNPs for Cigarettes per day.*

|  | | **MR – total effect** | | | | | **MVMR – direct effect** | | | | |
| --- | --- | --- | --- | --- | --- | --- | --- | --- | --- | --- | --- |
| **Exposure** | **nSNPs** | **OR** | | **95% C.I.** | | **P-value** | **OR** | | **95% C.I.** | **P-value** | |
| *Smoking Initiation* | |  | |  | |  |  | |  |  | |
| age_10 | 265 | 1.22 | | [1.13 1.32] | | 2.35E-06 | 0.97 | | [0.86 1.09] | 0.614 | |
| adult | 467 | 1.36 | | [1.26 1.47] | | 1.77E-16 | 1.40 | | [1.27 1.55] | 3.62E-11 | |
| *Smoking Cessation* | |  | |  | |  |  | |  |  | |
| age_10 | 267 | 1.12 | | [1.02 1.24] | | 0.007 | 0.95 | | [0.85 1.07] | 0.447 | |
| adult | 468 | 1.23 | | [1.14 1.33] | | 4.76E-07 | 1.30 | | [1.15 1.46] | 4.89E-06 | |
|  | **nSNPs** | | $\boldsymbol{\beta}$ | **Std. Err** | **P-value** | | | $\boldsymbol{\beta}$ | **Std. Err** | | **P-value** |
| *Cigarettes per day* | | |  |  |  | | |  |  | |  |
| age_10 | 265 | | 0.11 | [0.05 0.17] | 6.80E-06 | | | -0.04 | [-0.10 0.02] | | 0.252 |
| adult | 462 | | 0.23 | [0.19 0.27] | 6.76E-27 | | | 0.26 | [0.20 0.32] | | 1.34E-18 |

nSNPs; number of SNPs associated with the exposure, OR; MR estimated odds ratio for binary outcomes, $\beta$; MR effect estimate for continuous outcome, 95% CI; 95% Confidence Interval for MR estimate, P-value; P-value for MR estimate.
